# Time-dependent LSTM for Survival Prediction and Patient Subtyping in Kidney Disease Trajectory

**DOI:** 10.1101/2024.09.25.24314409

**Authors:** Pumeng Shi, Chunmei Fu

## Abstract

Chronic kidney disease (CKD) affects over 10% of the global population and is projected to become the fifth leading cause of years of life lost (YLL) by 2040. Accurate prediction of CKD progression to end-stage kidney failure (ESKF) is critical for timely interventions that can slow or halt disease progression. However, current models often fail to address the complexities of time-varying biomarkers like estimated glomerular filtration rate (eGFR) and the irregular nature of longitudinal health data, resulting in suboptimal predictions. In this study, we develop a Time-dependent Long Short-Term Memory (TdLSTM) network to analyze longitudinal eGFR data and predict time-to-ESKF. Our model is specifically designed to handle irregular time intervals and temporal dynamics, capturing nuanced patterns of CKD progression. We conducted experiments on two independent CKD cohorts, MASTERPLAN and NephroTest, using patient data including age, gender, eGFR, UACR, and diagnosis. The TdLSTM model outperformed traditional and state-of-the-art predictive models, demonstrating superior accuracy in estimating time-to-ESKF and identifying subtypes of CKD progression through unsupervised clustering. By leveraging the temporal dynamics of biomarkers, our approach offers a robust tool for personalized survival prediction and risk stratification. These findings highlight the potential of deep learning in improving CKD management and identifying high-risk patients in time for effective intervention.

## 1. Introduction

Chronic kidney disease (CKD) affects over 10% of the global population, impacting more than 850 million individuals worldwide (Francis et al., 2024). By 2040, CKD is expected to rank as the fifth leading cause of years of life lost (YLL) globally (Foreman et al., 2024). Individuals with CKD face a heightened risk of progression to end-stage kidney failure (ESKF), as well as increased rates of hospitalization and cardiovascular mortality (Al Alawi et al., 2017). Early detection of CKD is critical, as timely intervention can significantly slow or halt disease progression. Preventing kidney function decline is particularly effective in patients with mild to moderate CKD who are at high risk of advancing to ESKF. Thus, accurately predicting CKD progression holds immense clinical importance. However, previous studies have often focused on overall disease progression, neglecting individualized risks of transitioning to ESKF and failing to address censored data or capture personalized temporal variations. Uncovering patterns in kidney disease trajectories, especially within sparse and irregular longitudinal data, is essential for a more nuanced understanding of CKD development.

We focuse on identifying potential patterns while forecasting the occurrence of end-stage kidney failure (ESKF) in a cohort of CKD patients. Specifically, we aim to predict the time-to-ESKF by analyzing comprehensive biomarkers. Among these, the estimated glomerular filtration rate (eGFR) – a key biomarker for CKD – measures kidney function and determines disease stage (Coresh et al., 2014). In this study, ESKF is defined as dialysis initiation, preemptive transplantation, death due to kidney failure, or an eGFR below 15 ml/min/1.73m2. The eGFR is calculated using blood creatinine levels alongside factors such as age, body size, and gender. A low eGFR indicates impaired kidney function, with lower values correlating to an elevated risk of CKD progression to kidney failure. Additionally, reduced eGFR is strongly associated with adverse cardiovascular and renal outcomes, with risks increasing as eGFR declines. To address this, we need to estimate eGFR values over time up to a specified landmark. Importantly, in this longitudinal study, eGFR is a time-varying variable within a two-year prediction horizon, adding complexity to prediction efforts. Improper handling of time-varying eGFR can result in underestimation of standard errors and overly low p-values, skewing the association between eGFR dynamics and CKD progression.

However, current models are inadequate for addressing time-varying biomarkers like eGFR, as they fail to capture its evolution, highlighting a critical gap that this study seeks to address. The duration between consecutive elements in a sequence of time-varying biomarkers is often irregular, especially in health data such as the CKD data (which will be discussed below), where patient records are unstructured, with follow-up intervals ranging from days to months or even years. These varying time gaps can signal different health conditions; for example, frequent admissions might indicate a deteriorating health status, while long intervals between records may reduce the relevance of past data for predicting current outcomes. Traditional LSTM architectures do not account for such time irregularities in longitudinal data. In healthcare, where the timing between hospital visits or clinical measurements is crucial for decision-making, an LSTM architecture that incorporates these irregular elapsed times is essential for accurately analyzing temporal data.

In this paper, we develop a deep learning model that effectively handles time-varying variables to estimate the impact of eGFR decline on prognosis and explore subtypes of patients with different progression rates while performing survival prediction. We design Time-dependent Long Short-Term Memory (TdLSTM) networks, which are well-suited for processing time-varying inputs by storing and updating context information. Unlike feed-forward neural networks, LSTM – an advanced form of recurrent neural networks (RNNs) – uses internal memory to process sequential data (Zhu et al., 2023a). The feedback loop within TdLSTM units allows the network to retain information from past inputs indefinitely, enabling it to exhibit temporal dynamic behavior (Baytas et al., 2017). Moreover, TdLSTMs excel in managing long-term dependencies through a gated architecture that determines what information to retain or discard from previous time steps. This capability overcomes the limitations of traditional RNNs, making LSTMs ideal for analyzing the progression of CKD over time when taking the underlying previous health status into account.

We conducted experiments on two cohorts of moderate CKD patients characterized by five variables: age, gender, eGFR, UACR, and diagnosis. Patients with an eGFR >30 ml/min/1.73m2 were extracted from the MASTERPLAN and NephroTest studies. To predict eGFR and time-to-ESKF, we trained the model on the MASTERPLAN dataset and externally validated its performance on the NephroTest dataset for both baseline and follow-up studies. The TdLSTM model, with cross-validation on the combined datasets, was evaluated against other predictive models. Additionally, unsupervised clustering was used to identify subtypes associated with CKD progression. To comprehensively assess predictions, we employed three evaluation metrics to measure model performance. By examining the errors in estimated time-to-ESKF, we demonstrated the effectiveness of our deep learning model. A key advantage of our approach lies in its ability to learn representative embeddings that capture evolutionary clinical information, enabling detection of temporal differences in disease progression among individuals through unsupervised clustering. The experimental results on the two kidney disease cohorts (MASTRPLAN and NephroTest) and the comparison between TdLSTM and other state-of-the-art models demonstrate that our predictive model performs the best in both baseline and longitudinal studies and the promising of our approach in identifying CKD subtypes and those patients at high risk in time.

## 2. Related Work

This paper focuses on longitudinal survival prediction in chronic kidney disease (CKD). Accordingly, we will review existing research on longitudinal studies utilizing machine learning and survival models, as well as studies related to CKD prediction.

### 2.1. Time-varying Survival Prediction

Survival analysis has traditionally been dominated by the semi-parametric Cox proportional-hazards model (Cox, 1972), favored for its ease of use, proven effectiveness, and interpretability. However, the growing availability of diverse and complex datasets, such as those involving time-dependent covariates, has introduced challenges to conventional statistical approaches, driving significant interest in integrating machine learning techniques with survival models. For instance, Zhang et al. (2020b, 2019) developed a time-dependent survival neural network that estimates latent failure risks additively and performs multiple binary classifications to predict survival probabilities. Specifically, Zhang et al. (2020b) proposed a feed-forward architecture for such a network, leveraging repeated measures to estimate survival risks dynamically. Additionally, Lee et al. (2018) employed a deep neural network to directly learn the distribution of survival times, enabling the examination of covariate-risk relationships over time without relying on assumptions about the stochastic process.

Beyond neural networks, other machine learning techniques, including multi-task learning, Gaussian processes, Bayesian inference, active learning, transfer learning, and feature engineering, have been applied to survival analysis. For instance, the effects of variables often change over time with longer follow-up periods (Fisher and Lin, 1999), necessitating methods that accommodate time-varying coefficients instead of static ones. In this context, **?** introduced the multi-task logistic regression (MTLR) model, which directly learns patient-specific survival distributions by combining multiple local logistic regression models in a dependent manner.

### 2.2. Machine Learning for CKD Prediction

In chronic kidney disease (CKD) research, machine learning approaches have been widely adopted to improve prediction and diagnosis. Zisser and Aran (2024) proposed a transformer-based architecture to predict the survival probability of 130,000 CKD stage 3 patients over a 48-month horizon. Similarly, Tan et al. (2023) developed a supervised long short-term memory (LSTM) autoencoder to identify and analyze acute kidney injury (AKI) subphenotypes with prognostic and therapeutic implications, using K-means clustering for subphenotype identification. Xu et al. (2022) introduced a multivariate time-series prediction model based on recurrent neural networks (RNNs) for dynamic AKI prediction in hospitalized patients, incorporating variables such as demographics, admission details, lab investigations, and prescriptions.

Other studies focused on large-scale applications. For instance, Wu et al. (2023) utilized a nationwide multicenter dataset from China encompassing over 7 million hospitalized patients to dynamically predict in-hospital death and dialysis in AKI patients. Meanwhile, Bai et al. (2022) investigated machine learning methods for predicting end-stage kidney disease (ESKD) risk in CKD patients over a 5-year period. Subashini and Venkatesh (2023) developed a deep learning framework to enhance CKD diagnosis using imbalanced medical datasets, while Marthin and Tutkun (2023) introduced an attention mechanism, Risk Information Weights (RIW), alongside an external autoencoder (ExternalAE) for feature selection, to extract complex characteristics linked to cause-specific clinical events.

Innovative approaches for CKD forecasting and subtype identification have also emerged. For example, Srivastava et al. (2022) applied preprocessing, data aggregation, and classification techniques to automate CKD severity detection using UCI repository data. Dashtban et al. (2023) used seven unsupervised machine learning methods, including K-means and Clustering Large Applications, to identify CKD subtypes and evaluate their prognostic validity for 5-year mortality and hospital admissions. Additionally, Joo et al. (2023) proposed a predictive CKD risk score using deep learning applied to retinal photographs, validated on longitudinal cohorts from the UK Biobank and the Korean Diabetic Cohort. The resulting Reti-CKD score outperformed traditional eGFR-based methods in stratifying future CKD risk.

Further advancements include Chowdhury et al. (2024), who developed a heterogeneous ensemble model using 11 machine learning algorithms and 22 features from type 1 diabetes (T1D) patients to predict 10-year CKD risk, and Khalid et al. (2024), who evaluated machine learning techniques for predicting CKD progression, focusing on diabetic kidney disease, proteinuria, and eGFR decline. Zhu et al. (2023b) utilized longitudinal electronic health records (EHRs) to build an RNN model predicting CKD progression from stages 2 and 3 to stages 4 and 5, highlighting the importance of time-series data like eGFR measurements. Lastly, Zheng et al. (2024) classified patients into progression and non-progression groups using Cox models and random survival forests, while Liu et al. (2024) employed a super learner strategy to predict kidney failure and mortality, incorporating the kidney failure risk equation and accounting for competing mortality risks.

## 3. Proposed Method

To overcome the limitations of LSTM in handling irregular time intervals in longitudinal data, we propose a novel architecture – time-dependent LSTM (TdLSTM). This approach incorporates the varying elapsed time between consecutive elements in a sequence as part of the patient’s medical history, enabling the model to dynamically adjust its memory content. The study design, illustrated in Figure 1, involves training TdLSTM models on CKD data while preserving temporal information through a backpropagation learning process. The TdLSTM encoder generates variable representations that facilitate clustering patients into distinct subtypes with differing progression rates. Meanwhile, the TdLSTM decoder produces embeddings of patient variables for both eGFR prediction and time-to-ESKF prediction. To enhance accuracy, we also developed a survival time estimation method to predict the exact time-to-ESKF based on the modeled survival probability.

**Figure 1.**
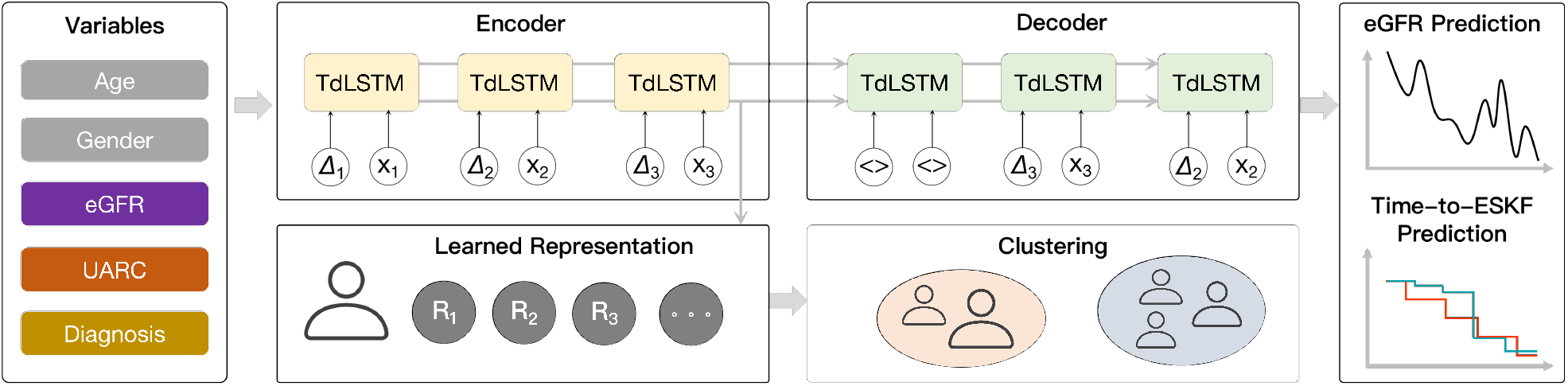
An overview of the study design. Patients’ clinical data, including demographics (age and gender), eGFR, UARC, and diagnosis. The proposed TdLSTM model encoder generates variable representation for clustering patients and the decoder learned the embedding is used for eGFR and time-to-ESKF prediction.

**Figure 2.**
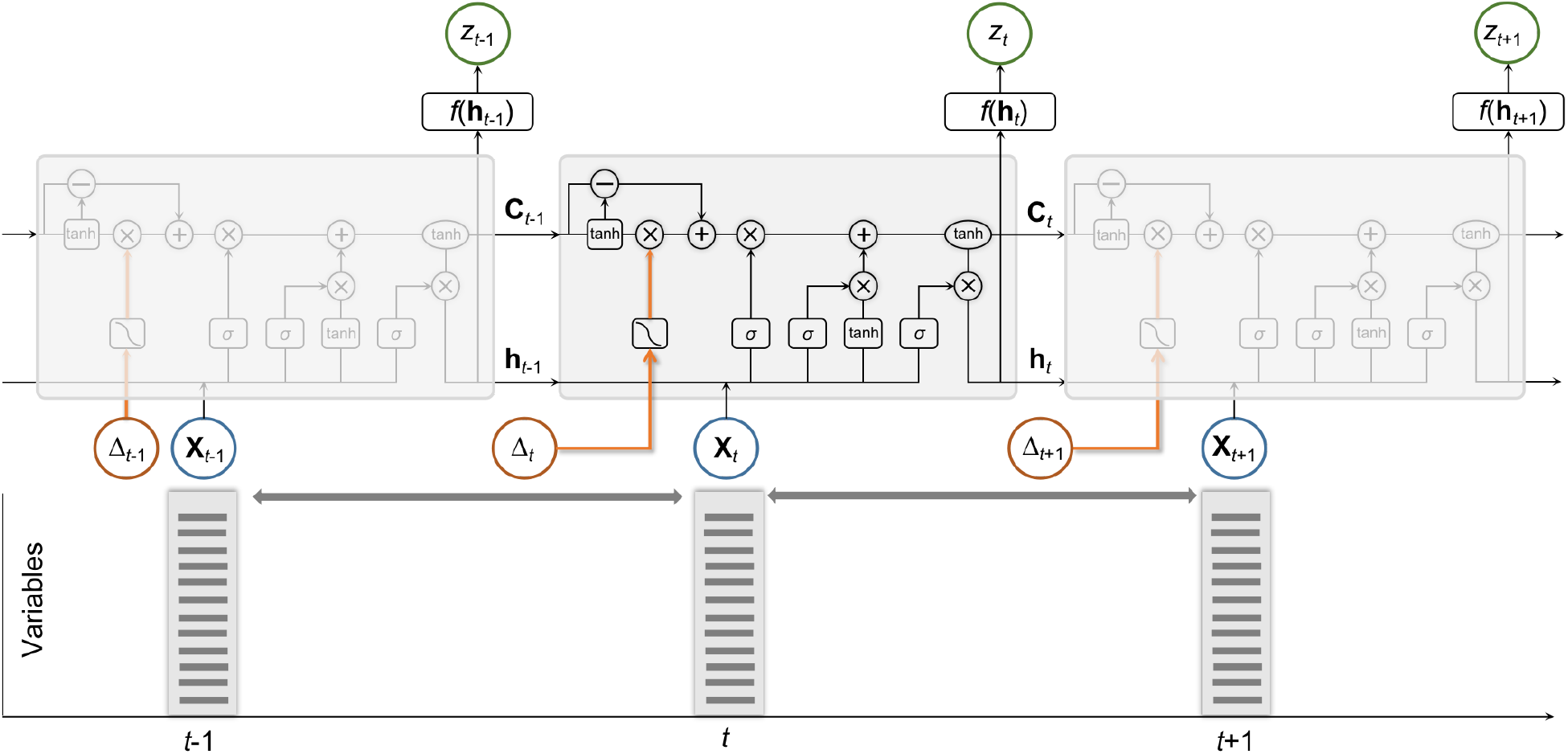
Illustration of the proposed time-dependent LSTM unit, and its application on analyzing our kidney patients’ health records. Green boxes indicate networks and yellow circles denote point-wise operations. TD-LSTM takes two inputs, input record and the elapsed time.

### 3.1. Data Formalization

Given *N* patients described by *V* variables, **X** = (**x**_1_, **x**_2_,…, **x**_*N*_) ∈ ℝ^*N*×*V*^, the eGFR prediction model outputs a series of eGFR values *Z* = (*z*_1_, *z*_2_, ⋯), while the time-to-ESKF prediction model generates an outcome vector of survival probabilities *P* = (*p*(*t*_1_),*p*(*t*_2_),⋯) over the disjoint time points *t*_1_,*t*_2_,⋯ for these patients. In our eGFR learning process, the outcomes *Z* can be modeled as a function of the variables **X**, where the learning objective is to find the mapping *f* such that **Z** = *f* (**X**). The survival curves can be plotted by identifying a survival function *S* that yields the survival probability *p* = *S*(*t*) for all *t* = *t*_1_, *t*_2_, ⋯. In time-to-ESKF prediction, we denote the ESKF response variables by (𝒯, ζ), where 𝒯 is the measurement time, *i*.*e*., the minimum of ESKF time *T* and (right-)censoring time *C, i*.*e*.,

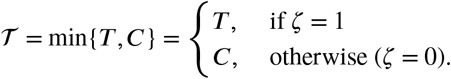

The indicator ζ = 𝟙 {*T* ≤ *C*} takes the value 1 if the patient reached end-stage kidney failure (ESKF) and 0 otherwise. In other words, ζ ∈ {0, 1} represents whether the event of ESKF occurrence has been observed or censored. We assume, without loss of generality, that the patients are ordered in ascending order of observation time. Additionally, *T* (the event time) and *C* (the censoring time) are assumed to be conditionally independent given the observed data. This implies that the time until ESKF (*T*) and the censoring time (*C*) do not influence each other once the covariates are accounted for.

### 3.2. Time-dependent LSTM Model

The schematic of the proposed TdLSTM includes LSTM units, each comprising an input, three gates (input, forget, and output), and an output activation function. The output of the unit is recurrently connected back to the unit input and all of the gates, *i*.*e*., **H**_*t*_ = **O**_*t*_ ⊙ tanh(**C**_*t*_), where ⊙ is a Hadamard (element-wise) product (Zhang et al., 2020a), and the output gate **O**_*t*_ modulates the amount of memory content exposure. Unlike a simple recurrent unit that computes a weighted sum of the input variables and applies a nonlinear function, each LSTM unit maintains a memory **C**_*t*_ at time *t*. This memory cell is updated by partially forgetting the adjusted previous memory 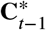 and adding new candidate memory content 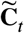 such as 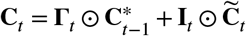.

- This decomposition is data-driven, with the parameters of the decomposition network being learned simultaneously alongside the other network parameters through back-propagation. There is no strict requirement for the type of activation function used in the decomposition network, though the tanh function has been shown to perform slightly better, where 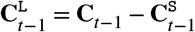. The short-term memory 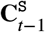 is obtained by a network, *i*.*e*., 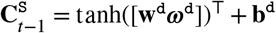. After obtaining the short-term memory, it is adjusted by applying an elapsed time weight, resulting in the discounted short-term memory, *i*.*e*., 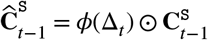. This adjustment accounts for the varying time intervals between consecutive elements in the sequence, ensuring that the memory reflects the temporal dynamics of the data more accurately. As a guideline, let Δ_*t*_ represent the elapsed time between **x**_*t*−1_ and **x**_*t*_. The function *ϕ*(⋅) is a heuristic decaying function that adjusts the influence of the short-term memory based on Δ_*t*_. The function *ϕ*(⋅) should be monotonically non-increasing, meaning that as Δ_*t*_ increases, the effect of the short-term memory decreases. Different types of monotonically non-increasing functions can be chosen for *ϕ*(⋅) depending on the measurement type of the durations specific to the application domain. In healthcare, where the elapsed time between successive records can range from days to years, we normalize the elapsed time to a common unit, such as days. For datasets with large elapsed times, we might use 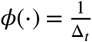 or 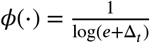, ensuring that the short-term memory’s influence diminishes appropriately with increasing elapsed time. Finally, to reconstruct the adjusted previous memory 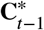, the complement subspace of the long-term memory 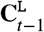 is combined with the discounted short-term memory 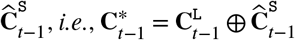. Here, ⊕ denotes the combination of the complement subspace of the long-term memory and the discounted short-term memory.
- The input weights in terms of input gate, forget gate, output gate and cell unit in hidden layer are denoted by **w**^i^,**w**^f^,**w**^o^,**w**^c^ ∈ ℝ^*D*×*V*^ respectively, the recurrent weights by *ω*^i^,*ω*^f^,*ω*^o^,*ω*^c^ ∈ ℝ^*D*×*D*^, and the bias as **b**^i^,**b**^f^,**b**^o^,**b**^c^ ∈ ℝ^*D*^. The input gate **I**^*τ*^, forget gate Γ^*τ*^, output gate **O**^*τ*^, and candidate cell 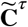 are computed by

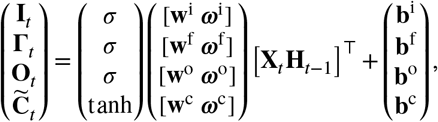

where [**X**_*t*_ **H**_*t*−1_]^⊤^ is the concatenation of the two vectors: input variables **X**_*t*_ and **H**_*t*−1_.

### 3.3. Survival Learning

Our model’s learning process encompasses estimating survival probabilities, predicting survival times, and identifying patient subtypes.

#### 3.3.1. Probability Estimate

For patient *i* (*i* = 1, 2,…, *N*) with variables **X** at time *t*, TdLSTM estimates the survival probability (*i*.*e*., the probability of staying before the end stage) as follows:

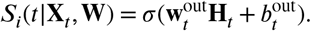

TdLSTM cannot be an effective prediction model unless it achieves the objective that *the predicted survival probabilities approach the actual progression of ESKF*. We leverage the Kullback-Leibler (KL) divergence between the distributions of *S*_*i*_(*t*|**X, W**) and survival status ζ_*i*_(*t*) ∈ {0, 1} to quantify such approachability, as follows:

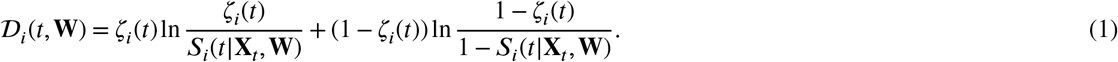

The optimal weights are designed to ensure that *S*_*i*_(*t*|**X**_*t*_, **W**) closely approximates 1 if patient *i* remains free of ESKF at time *t*_*k*_, and approaches 0 otherwise. Here, outputs of 1 and 0 correspond to definitively true and definitively false predictions, respectively. Our learning objective is to minimize 𝒟_*i*_(*t*, **W**) across the time points *t*_0_,*t*_1_,*t*_2_,…,*t*_*K*_, where survival statuses are available for all *N*_tr_ training patients. In this study, we set the time interval to one month, with *t*_0_ = 0, *t*_1_ = 1 month, and so on, up to *t*_*K*_ = 24 months.

It is important to note that the survival probability *S* naturally decreases from 1 to 0 over time, spanning the period from the initial state to the onset of ESKF. This behavior signifies that survival probability should decrease monotonically as time progresses. Consequently, the minimization process must adhere to this inherent monotonicity constraint. To address this, we employ a penalty method (Zhang et al., 2020b) that reformulates the constrained optimization problem into a series of unconstrained optimization problems. Specifically, we utilize the static penalty method (Michalewicz and Schoenauer, 1996) to penalize violations of the inequality constraints, ensuring that the survival probability *S* aligns with the expected monotonic behavior. The optimization problem is then expressed as minimizing the average error, computed as:

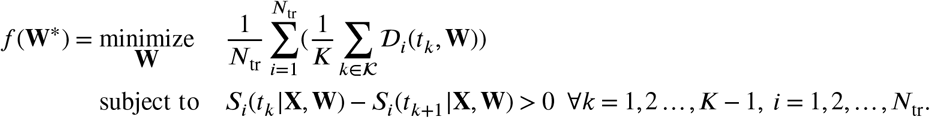

The penalty method, which converts a constrained minimization problem into a series of unconstrained optimization problems, has proven to be highly effective. The solution to the unconstrained problem converges to that of the original constrained problem. By leveraging the advantages of the exterior-point method (EPM) (Yamashita and Tanabe, 2010), we define the following objective function to account for violations of the survival probability constraints:

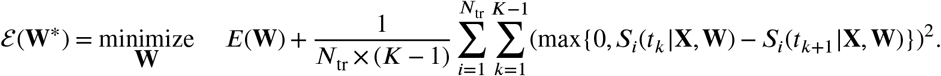

Model training is performed using the forward-only Levenberg-Marquardt algorithm (Wilamowski and Yu, 2010), which combines the computational efficiency of the Gauss-Newton algorithm (Gill and Murray, 1978) with the robustness of the steepest descent method.

#### 3.3.2. Time Estimate

After obtaining the predicted survival probabilities at various time points from our model, using the learned parameters 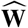, the next step involves estimating the time-to-ESKF for the test individual. To begin this process, we define the relative error (RE) at time *t* as follows:

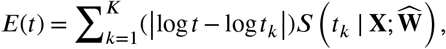

thereby seeking the time at which *E* is the lowest, *i*.*e*., the time-to-ESKF is

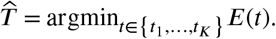

#### 3.3.3. Patient Subtyping

CKD patient subtyping is framed as an unsupervised clustering problem, given the lack of prior information about the groups within the patient cohort. To effectively cluster temporal and complex electronic health record (EHR) data, it is essential to have an efficient representation that captures the structure of patients’ temporal records. Autoencoders offer an unsupervised method to directly learn a mapping from the original data to a compressed representation. Specifically, LSTM autoencoders are well-suited for encoding longitudinal data. A single-layer autoencoder may require more iterations to minimize the reconstruction error, particularly when the learned representation is of lower dimensionality than the original input. Moreover, mapping to a lower-dimensional space introduces additional complexity, as more details of the high-dimensional input must be captured. Hence, we employ a two-layer time-dependent LSTM (TdLSTM) autoencoder mechanism to process a small sequence consisting of three elements. The hidden state and cell memory of the TdLSTM encoder at the end of the input sequence are used as the initial hidden state and memory content of the TdLSTM decoder. Initially, the input element and the elapsed time for the decoder are set to zero, with the first output being the reconstruction of the last element of the original sequence. Once the reconstruction error 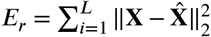 is minimized, the TdLSTM encoder is applied to the original sequence to obtain the learned representation, which corresponds to the hidden state of the encoder at the end of the sequence. This learned representation serves as a compact summary of the patient’s temporal data, enabling effective subtyping and facilitating further analysis.

## 4. Experiments

In this section, we will present the data in detail and the experimental setup, and analyze the experimental results.

### 4.1. Data Statistics

We collected data from the MASTERPLAN (van Zuilen et al., 2012) and NephroTest (Froissart et al., 2005) studies, where each patient was recorded with five variables: age, gender, eGFR, urine albumin/creatinine ratio (UACR), and diagnosed nephropathy (including diabetic nephropathy, glomerulonephritis, hypertension or vascular disease, polycystic kidney disease, tubulointerstitial nephritis, and other or unknown). The MASTERPLAN study is a randomized controlled trial comparing intensified care, facilitated by a nurse practitioner, to standard care provided by a nephrologist. Serum creatinine levels were standardized to a central laboratory in the MASTERPLAN study. In the NephroTest study, plasma creatinine was initially measured using a modified kinetic Jaffe colorimetric method with a Konelab 20 analyzer (Thermo-Fisher Scientific) from the study’s inception until 2008, and later with an enzymatic assay. Creatinine values obtained with the Jaffe method were standardized to an IDMS-traceable creatinine assay as previously described (Moranne et al., 2009). We calculated eGFR using the CKD-EPI equation for serum creatinine and selected 505 patients at stage 3 with eGFR >30 ml/min/1.73m2 from the MASTERPLAN study and 398 patients from the NephroTest study (Levey et al., 2009). During the follow-up period, 11% (*n* = 63) of the MASTERPLAN patients reached ESKF, while in NephroTest were 4% (*n* = 14) reached ESKF. Compared to the MASTERPLAN, eGFR measurements in NephroTest were less frequent due to the annual follow-up schedule. Both studies received ethical approval from the respective committee (local IRB UMC Utrecht and CCTIRS MG/CP09.503 France), and all participants provided written informed consent. The time-to-ESKF is calculated from a 2-year landmark point up to a 4-year total follow-up period. The prediction horizon represents the maximum duration for which the models assess ESKF risk, while the landmark signifies the time since baseline when patient data is included in the model. Follow-up was censored upon reaching ESKF, death, or at the end of the 4-year period, whichever occurred first.

### 4.2. Statistics

The average age of patients in the MASTERPLAN and NephroTest cohorts was 58 and 56 years, respectively. The gender distribution was similar across both cohorts, with 69% male participants in MASTERPLAN and 67% in NephroTest. Regarding nephropathy diagnoses, diabetic nephropathy remained stable at 9-10% in both cohorts. Glomerular nephropathy was observed in 18% of MASTERPLAN participants and 19% of NephroTest participants. Vascular nephropathy was more prevalent in MASTERPLAN (27%) compared to NephroTest (25%). Congential nephropathy showed a large decrease in NephroTest (7%) compared to MASTERPLAN (12%). Interstitial nephropathy remained stable at 11% in MASTERPLAN and 12% in NephroTest. The category of other or unknown nephropathy was more common in the NephroTest cohort, while in the MASTERPLAN cohort, it remained around 22%. The estimated glomerular filtration rate (eGFR) was also comparable between the two cohorts. Baseline eGFR values were around 50 ml/min/1.73m2 in MASTERPLAN and 52 ml/min in NephroTest, both declining to 48-49 ml/min/1.73m2 in longitudinal study. The urine albumin-to-creatinine ratio (UACR) showed same trends: in MASTERPLAN, UACR mostly decreased from 40mg/g (median) at baseline to 36mg/g (median) at longitudinal base, while in NephroTest, it decreased from 50 mg/g to 44mg/g. (Due to missing UACR values in longtidual MASTERPLAN study, the mean of UACR had a huge difference compared to others.) These detailed data provide insights into the stability and shifts in key health indicators and diagnoses within and between these two cohorts over time.

### 4.3. Setup

We compared the proposed method with the following state-of-the-art predictive models: TdCox (Fisher and Lin, 1999) extends the Cox model (Cox, 1972) to time-dependent covariates and has a survival function *S*(*t*) = *S*_base_ exp(***Β*x**^*t*^) with the baseline probability *S*_base_ when **x**^*t*^ = (0,0,…,0) and the regression coefficients ***Β*** describing how the survival probability responds to the covariates. RSF (Ishwaran et al., 2008) estimates conditional cumulative failure hazard by aggregating tree-based Nelson-Aalen estimators. MTLR (Yu et al., 2011) models survival probabilities for individuals with the event and censored individuals. The logistic regression coefficients are time-varying. DeepHit (Lee et al., 2018) employed a network architecture that consists of a single shared sub-network and a family of cause-specific sub-networks. We train the network by using a loss function that exploits both survival times and relative risks.

We implemented the models using PyTorch (version 2.4) and Scikit-learn (version 1.5.1). Missing data were addressed using multivariate imputation by chained equations (MICE) (Van Buuren and Oudshoorn, 2000) with the Scikit-learn “IterativeImputer” method, where a random forest algorithm was applied to perform imputation. Imputation was carried out five times, with the average of these results used as the final imputation.

### 4.4. Model Evaluation

To evaluate the model’s performance on the *N* patients, we redefine three independent metrics: the kidney failure AUC, the time-dependent concordance index (C-index), and the mean squared error (MSE), redefined as follows (1 is the indicator function)

- AUC provides a probability measure of classification ability at a pre-specified time snapshot (*e*.*g*., at *t*_*K*_ = 2 years in our case). It quantifies the model’s ability to address the issue *“Is patient i likely to reach ESKF by time t?”*.

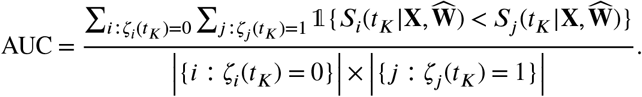
- As a generalization of the AUC, C-index estimates how accurately the model can answer the question *“Which of patient i and patient j is more likely to reach the ESKF?”*.

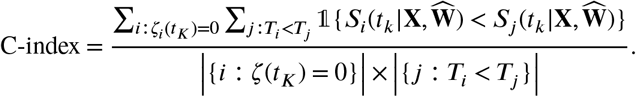
- MSE is the variant of Brier score that measures a prediction error across the data, *i*.*e*., the power of a model to address the question *“How accurate is the prediction that patient i will reach ESKF?”*.

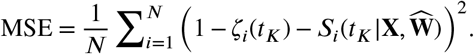

### 4.5. Subtyping Results

We clustered the patients into two distinct subgroups, each corresponding to a different progression pattern. In the MASTERPLAN study, 359 patients were assigned to the first subgroup (pattern 1), with 45 of these patients progressing from Stage 3 to ESKF, while 314 were censored. The second subgroup (pattern 2) included 146 patients, of whom 18 converted from Stage 3 to ESKF, and 128 were censored. In the NephroTest set, 281 patients were categorized into pattern 1, with 11 patients converting from Stage 3 to ESKF, and 270 being censored. Pattern 2 comprised 117 patients, of whom 2 progressed to ESKF, while 115 were censored. Figure 3 provides a visualization of these patterns using 2D t-SNE (t-distributed Stochastic Neighbor Embedding) (Van der Maaten and Hinton, 2008). Initially, we observed indistinct clusters when visualizing the baseline variables, as shown in subfigure A for MASTERPLAN and subfigure C for NephroTest. In these subfigures, patients are distributed with significant overlap in the 2-dimensional space formed by the original 5 variables. However, when we visualized the latent representation learned from the hidden layer of our model, a clear separation between the two clusters emerged. In this new 2-dimensional space, which combines baseline variables with time-to-ESKF information, the patients in both datasets formed two distinct and separable clusters, effectively distinguishing the two patterns (subtypes), as shown in subfigure B for MASTERPLAN and subfigure D for NephroTest.

**Figure 3.**
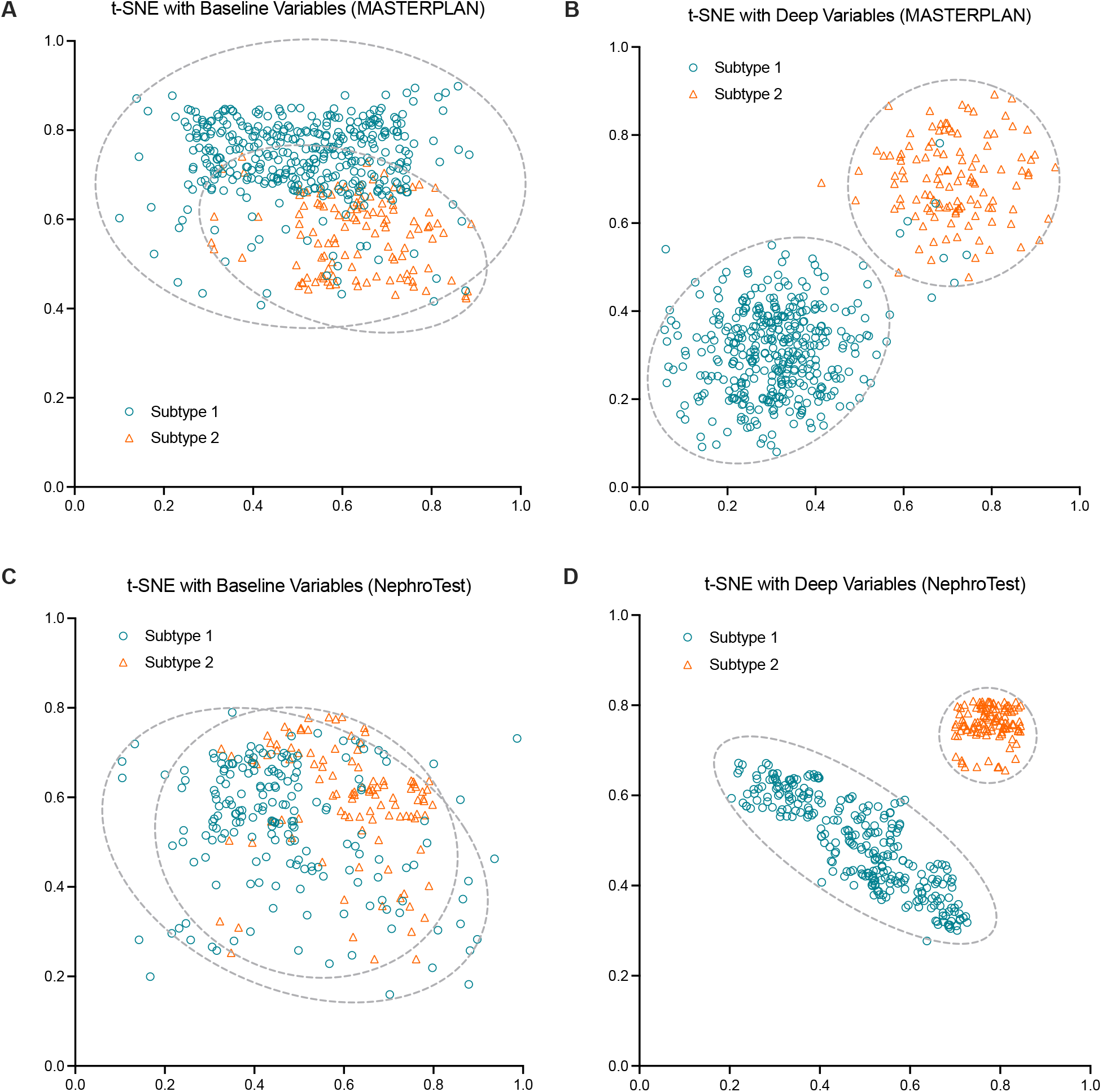
t-SNE for the distribution of the identified patterns on the MASTERPLAN and NephroTest datasets. Subfigures A and C are two-dimensional t-distributed stochastic neighbor embeddings (t-SNE) of the variables of the patients with two patterns on the MASTERPLAN and NephroTest follow-up datasets. Subfigures B and D are two-dimensional t-SNE by the hidden-layer activations of the model for the two patterns on the two follow-up datasets. The hidden-layer variables contain the representation of baseline variables and the temporal representation from the ESKF time prediction.

### 4.6. Time-to-ESKF Prediction

We compared our TdLSTM model against the Cox model using baseline eGFR, and the time-dependent Cox model incorporating time-varying eGFR, as well as the other four models. Figure 4 illustrates the calibration between predicted and observed risks of progression to ESKF at a 2-year landmark with a 2-year prediction horizon, resulting in a total follow-up of 4 years in the two cohorts. The competitors consistently overestimated or underestimated the risk of ESKF in both baseline and longitudinal analyses. The Cox model (Breslow, 1975) using only the baseline eGFR value demonstrated reasonable calibration among low-risk participants but tended to overestimate the risk in high-risk individuals. Conversely, the Cox model that included time-varying eGFR variables showed improved calibration, particularly in the longitudinal study. Our TdLSTM model achieved the best calibration overall, with predicted risks closely aligning with observed risks, especially in the longitudinal study. This demonstrates the superior ability of our model to provide accurate risk predictions over time, effectively capturing the dynamic nature of disease progression. We further assessed the performance of time-to-ESKF prediction by incorporating the discovered patterns as an independent variable, comparing it against the five baseline variables, using 5-fold cross-validation on the combined MASTERPLAN and NephroTest datasets. Figure 5 presents the average results from 25 cross-validation experiments, highlighting the significant impact of different patterns on the C-index, which measures the risk of progression for both baseline and follow-up studies. The time-to-ESKF prediction utilizing only the two identified patterns achieved a C-index of 0.62 (*p* = 0.17) for baseline data and 0.68 (*p* = 7.6e-16) for follow-up data. These results outperform the C-index values obtained using individual baseline variables: 0.56 for age, 0.51 for gender, 0.6 for eGFR, 0.57 for UACR, and 0.54 for diagnosis. Similarly, the pattern-based model outperformed the follow-up data results for the same variables, which were 0.58, 0.52, 0.64, and 0.6, respectively. This demonstrates the robustness of the discovered patterns in enhancing prediction accuracy compared to traditional baseline variables.

**Figure 4.**
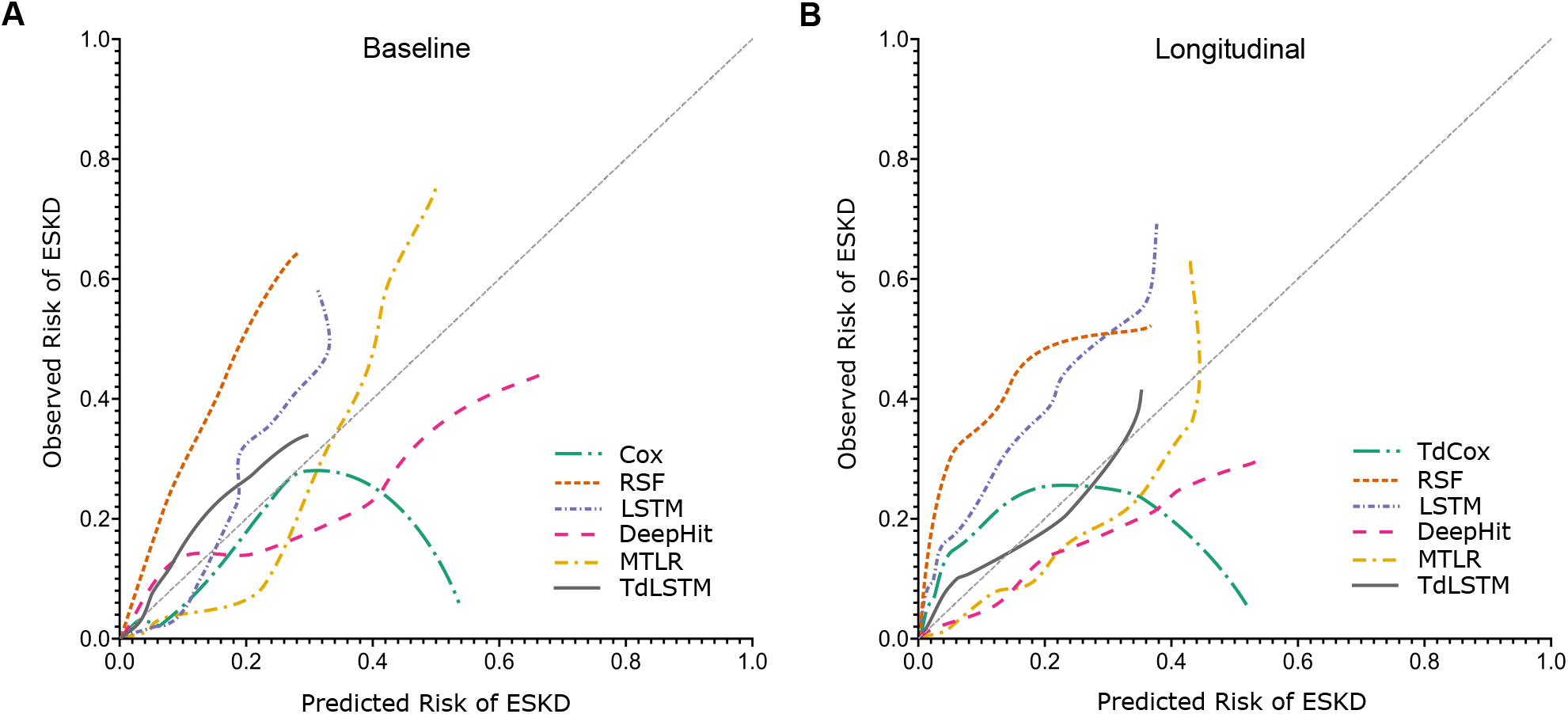
Calibration between predicted and observed risk of progression to ESKF at two years follow-up in the NephroTest cohort. The line indicates the average calibration line.

**Figure 5.**
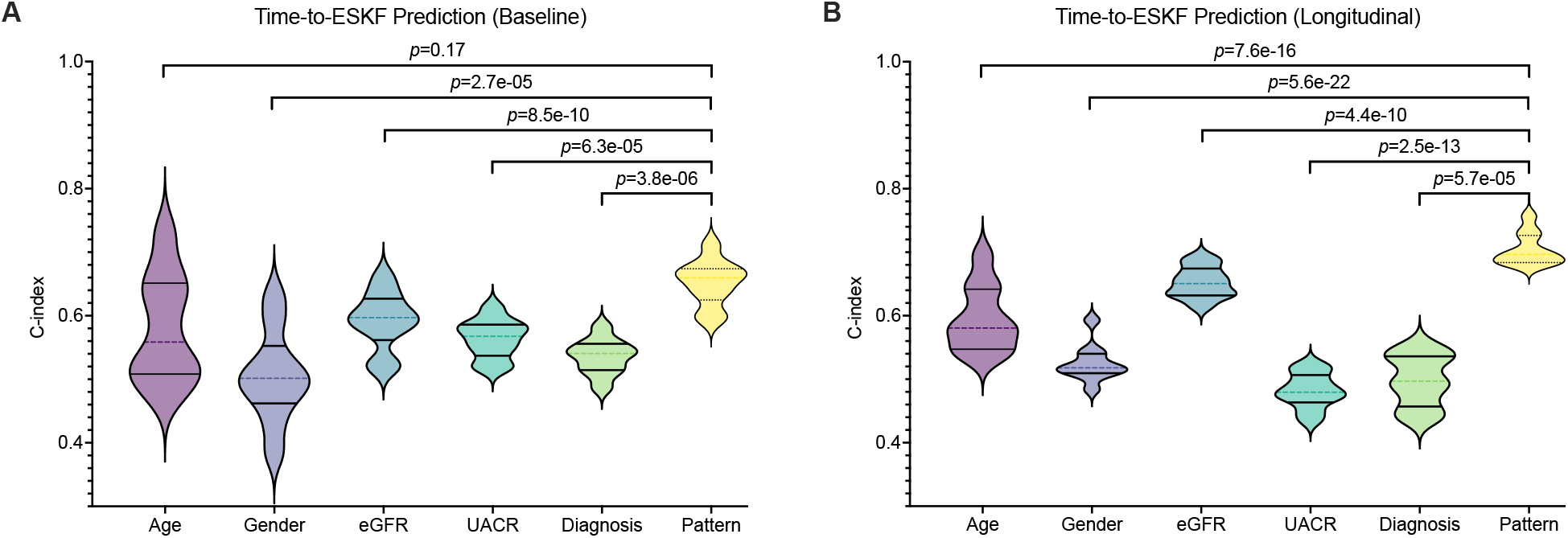
Predictive ability of the identified patterns. We evaluated the performance of time-to-ESKF prediction by C-index and compared the performance of the discovered patterns of the other five variables. The violin plots showed 5-fold cross-validation results on the mixed MASTERPLAN and NephroTest datasets for each variable.

To further assess the model’s effectiveness in predicting the probability of being free from ESKF, we compared the area under the ROC curve (AUC) associated with predicted probabilities over the 2-year follow-up period. Figure 6 illustrates the AUC results for 1-year and 2-year progression predictions based on baseline data (left) and follow-up data (right). The findings indicate that when using baseline variables, our model achieves an AUC of 0.833 for 1-year progression prediction and 0.707 for 2-year progression. However, when using follow-up variables, the AUC improves significantly, reaching 0.955 for 1-year progression and 0.84 for 2-year progression. This demonstrates the model’s enhanced predictive power when incorporating follow-up data, particularly in long-term predictions.

**Figure 6.**
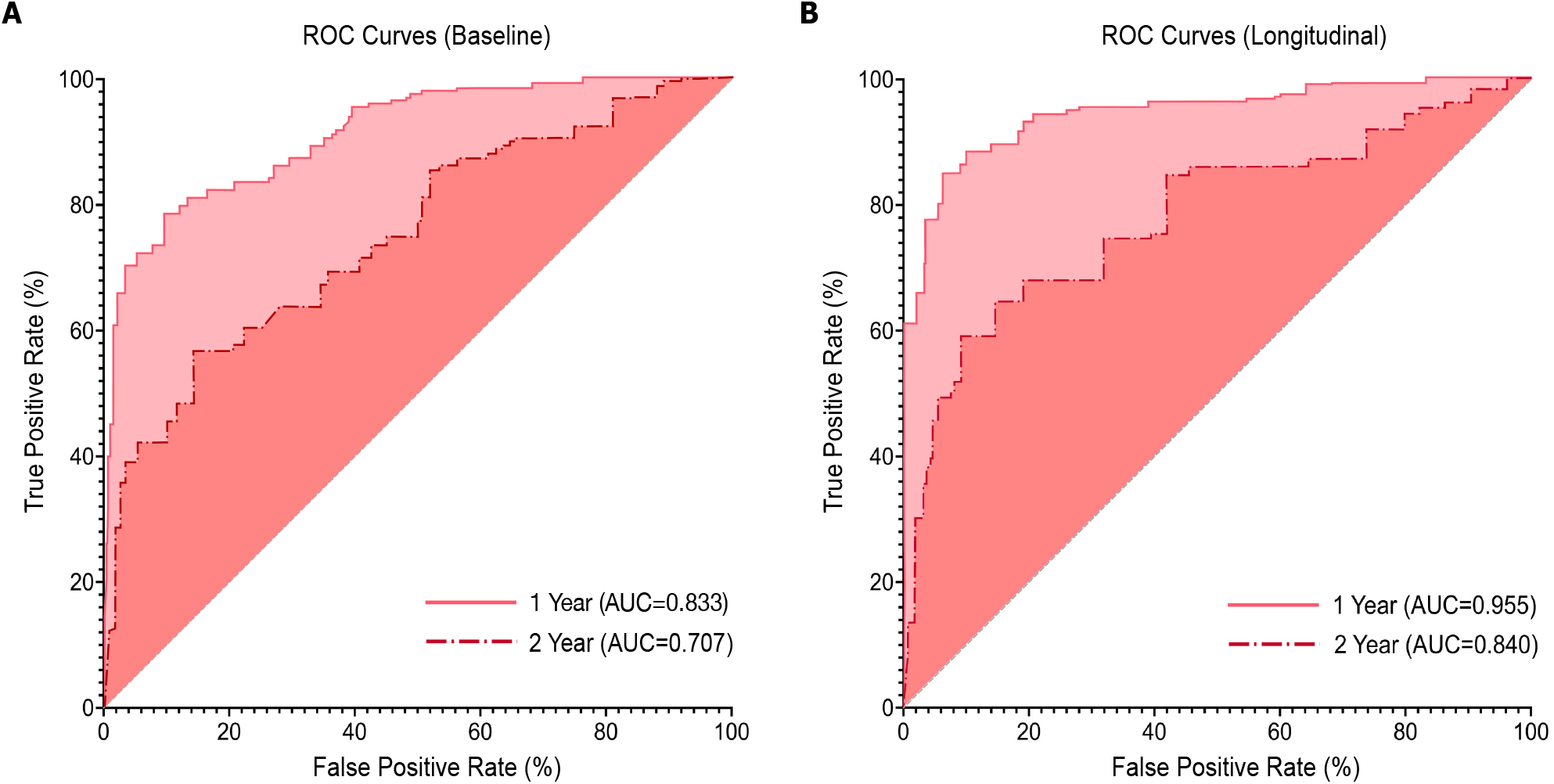
ROC curves for prediction of ESKF time on baseline and follow-up datasets, associated with 1 and 2 years of CKD progression.

We assessed the relative error of the estimated survival time, as summarized in Table 2. The proposed TdLSTM model significantly outperformed Cox-type models, yielding much lower relative errors (Yu et al., 2011). Specifically, TdLSTM achieved a relative error of 0.453 on baseline data (*p* = 3.9e-7) and 0.377 on follow-up data (*p* = 0.0002). These results highlight the superior accuracy of TdLSTM in estimating survival time, particularly when follow-up data is incorporated.

**Table 1.**
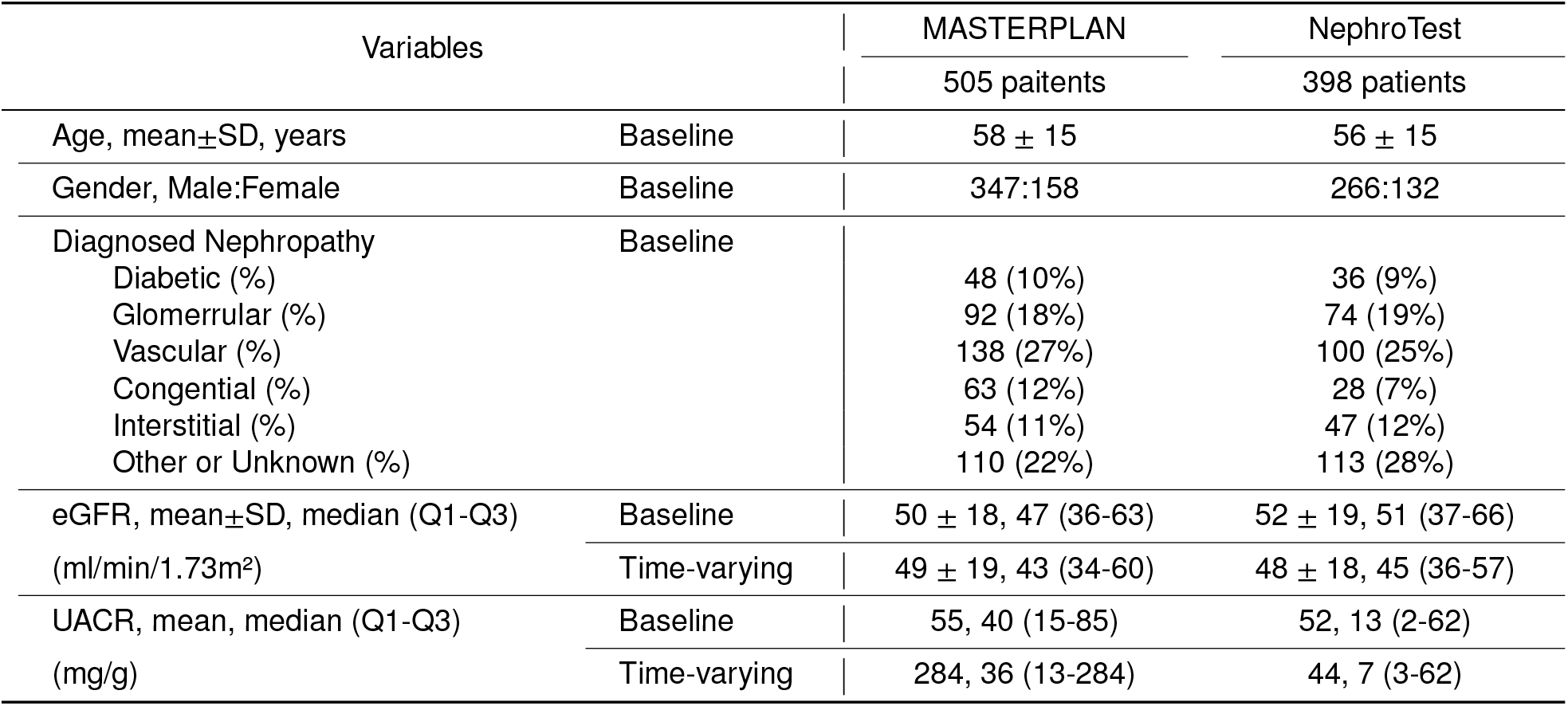
Characteristics and distribution of the MASTERPLAN and NephroTest cohorts.

| Variables |  | MASTERPLAN | NephroTest |
| --- | --- | --- | --- |
|  |  | 505 patients | 398 patients |
| Age, mean±SD, years | Baseline | 58 ± 15 | 56 ± 15 |
| Gender, Male:Female | Baseline | 347:158 | 266:132 |
| Diagnosed Nephropathy | Baseline |  |  |
| Diabetic (%) |  | 48 (10%) | 36 (9%) |
| Glomerular (%) |  | 92 (18%) | 74 (19%) |
| Vascular (%) |  | 138 (27%) | 100 (25%) |
| Congenital (%) |  | 63 (12%) | 28 (7%) |
| Interstitial (%) |  | 54 (11%) | 47 (12%) |
| Other or Unknown (%) |  | 110 (22%) | 113 (28%) |
| eGFR, mean±SD, median (Q1-Q3) | Baseline | 50 ± 18, 47 (36-63) | 52 ± 19, 51 (37-66) |
| (ml/min/1.73m<sup>2</sup>) | Time-varying | 49 ± 19, 43 (34-60) | 48 ± 18, 45 (36-57) |
| UACR, mean, median (Q1-Q3) | Baseline | 55, 40 (15-85) | 52, 13 (2-62) |
| (mg/g) | Time-varying | 284, 36 (13-284) | 44, 7 (3-62) |

**Table 2.**
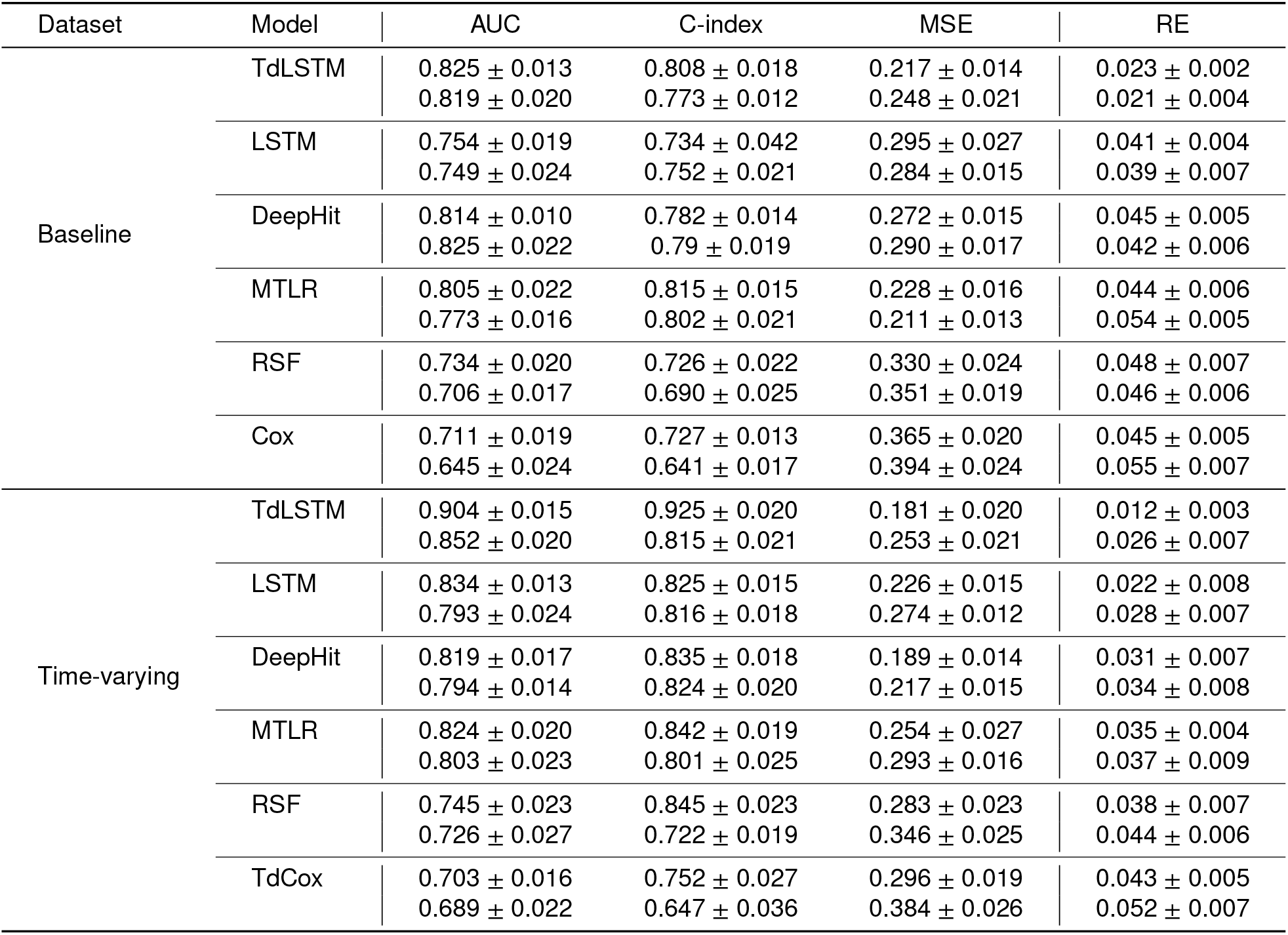
Comparison of the 5-fold cross-validation (5CV) results, and RE of the estimated survival times in the form of the mean ± standard deviation.

The results show that among the baseline models, TdLSTM delivers the most robust performance, achieving the highest AUC (0.825 ± 0.013) and C-index (0.808 ± 0.018), indicating strong discriminative ability and ranking accuracy. Its low MSE (0.217 ± 0.014) and RE (0.023 ± 0.002) reflect excellent survival time prediction accuracy and reliability. In comparison, LSTM demonstrates moderate effectiveness with slightly lower AUC (0.754 ± 0.019) and C-index (0.734 ± 0.042) and higher MSE (0.295 ± 0.027), indicating less accurate predictions. DeepHit and MTLR perform competitively, achieving relatively high AUC and C-index values, but their slightly elevated MSE and RE suggest room for improvement in survival time precision. RSF and Cox models consistently underperform, with lower AUC and C-index scores and significantly higher MSE and RE values, highlighting challenges in both discrimination and estimation accuracy. Overall, TdLSTM emerges as the most effective baseline model, combining superior ranking performance with reliable survival time estimates.

We compared the survival probability curves generated by competing models using a case study of a CKD patient who experienced failure within four months of follow-up. A survival curve was plotted based on the average failure-free survival probability predicted by each model. As shown in Figure 7, the TdLSTM model consistently predicts lower average probabilities across all follow-up periods compared to other models, particularly in longitudinal studies. This is primarily due to its ability to estimate latent risk effectively, thereby refining the relationship between latent risk and failure-free survival probability. Consequently, TdLSTM enables healthcare providers to issue warnings significantly earlier than other models, allowing for timely maintenance interventions that could help prevent potential failures.

**Figure 7.**
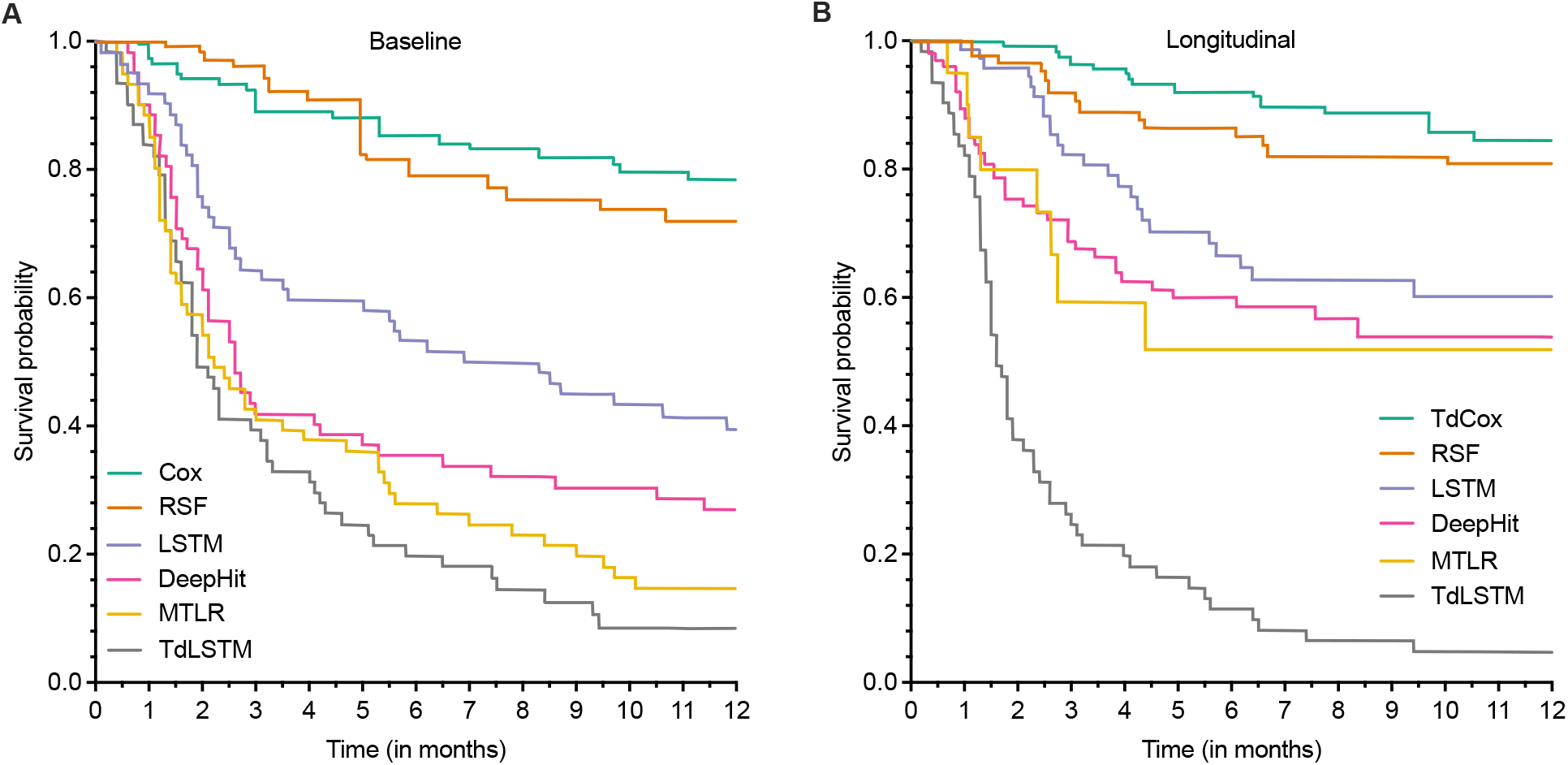
Change in survival probability curve for a patient that reached ESKF in 4 months of follow-up. Probabilities are predicted based on (A) baseline data and (B) longitudinal data.

### 4.7. Conclusion

his work presents a novel RNN-based model for predicting eGFR and time-to-ESKF in patients with chronic kidney disease (CKD). The model performs nonlinear prediction and survival analysis using longitudinal data, where each patient’s trajectory is represented as a latent state derived from baseline clinical variables and disease progression over time. Unlike traditional methods, this approach effectively handles time-varying variables and identifies distinct subtypes of CKD progression. To address long-term dependencies, we implemented an enhanced version of Long Short-Term Memory (LSTM) networks. While standard LSTMs assume uniformly spaced time intervals, our model adjusts for irregular time intervals, which is crucial for longitudinal data. This innovation enables the model to better capture the nonlinear progression of CKD, improving predictive performance over simpler methods that rely on linear concatenation. Our model preserves the temporal continuity of individual disease progression without explicitly modeling CKD stages. By aligning patients on the time axis and performing deep survival clustering, we incorporate both baseline variables and time-to-ESKF into the latent representations, revealing hidden patterns in patient trajectories. This results in a more nuanced understanding of disease progression, particularly in cases with heterogeneous trajectories.

However, the relatively small dataset limits the potential for extensive deep model parameter tuning. Failing to account for this uncertainty may lead to an underestimation of standard errors and an overly optimistic assessment of statistical significance (p-values), particularly when analyzing the association between time-varying exposures and eGFR slope, especially when comparing periods of progression and non-progression within the same patient.

## Data Availability

All data produced in the present study are available upon reasonable request to the authors

